# Chest computed tomography (CT) scan findings in patients with COVID-19: a systematic review and meta-analysis

**DOI:** 10.1101/2020.04.22.20075382

**Authors:** Mohammad Karimian, Milad Azami

**Author notes:** Correspondence: Faculty of Medicine, Ilam, Iran. E.

## Abstract

**Objectives:** Numerous cases of pneumonia of caused by coronavirus disease 2019 (COVID-19) were reported in Wuhan, China. Chest computed tomography (CT) scan is highly important in the diagnosis and follow-up of lung disease treatment. The present meta-analysis was performed to evaluate chest CT findings in COVID-19 patients.

**Materials and Methods:** All research steps were taken according to the MOOSE protocol and the final report was based on PRISMA guidelines. Each stage of the study was conducted by two independent authors. We searched the Web of Science, Ovid, Science Direct, Scopus, EMBASE, PubMed/Medline, Cochrane Library, EBSCO, CINAHL and Google scholar databases. The search was conducted on March 20, 2020. Grey literature was searched at medrxiv website. All analyses were performed using Comprehensive Meta-Analysis. The adapted Newcastle Ottawa Scale was used to evaluate the risk of bias. We registered this review at PROSPERO (registration number: CRD42019127858).

**Results:** Finally, 40 eligible studies with 4,183 patients with COVID-19 were used for meta-analysis. The rate of positive chest CT scan in patients with COVID-19 was 94.5% (95%CI: 91.7-96.3). Bilateral lung involvement, pure ground-glass opacity (GGO), mixed (GGO pulse consolidation or reticular), consolidation, reticular, and presence of nodule findings in chest CT scan of COVID-19 pneumonia patients were respectively estimated to be 79.1% (95% CI: 70.8- 85.5), 64.9% (95%CI: 54.1-74.4), 49.2% (95%CI: 35.7-62.8), 30.3% (95%CI: 19.6-43.6), 17.0% (95%CI: 3.9-50.9) and 16.6% (95%CI: 13.6-20.2). The distribution of lung lesions in patients with COVID-19 pneumonia was peripheral (70.0% [95%CI: 57.8-79.9]), central (3.9% [95%CI: 1.4-10.6]), and peripheral and central (31.1% [95%CI: 19.5-45.8]). The most common pulmonary lobes involved were right lower lobe (86.5% [95%CI: 57.7-96.8]) and left lower lobe (81.0% [95%CI: 50.5-94.7]).

**Conclusion:** Our study showed that chest CT scan has little weakness in diagnosis of COVID-19 combined to personal history, clinical symptoms, and initial laboratory findings, and may therefore serve as a standard method for diagnosis of COVID-19 based on its features and transformation rule, before initial RT-PCR screening.

## 1. Introduction

In December 2019, numerous cases of pneumonia of unknown cause were reported in Wuhan, Hubei Province, China. On January 7, 2020, the novel coronavirus, severe acute respiratory syndrome coronavirus-2 (SARS-CoV-2) was identified as a causative organism by Chinese experts by performing real-time polymerase chain reaction (real-time PCR) on patients’ respiratory tract specimens. It was subsequently named 2019-nCoV by World Health Organization (WHO) (1). There is also evidence that it can be transmitted through respiratory droplets and contact with infected patients as well as fecal–oral transmission (2, 3). Etiologically speaking, virulence of a pathogen may increase sharply during host shifts (4, 5), and in contrast, the virulence may decrease through prolonged host-parasite interactions (6).

Coronavirus Disease 2019 (COVID-19) is primarily transmitted through respiratory droplets and close contact, and the incubation period is usually between 1 and 14 days. The common symptoms include fever, dry cough, fatigue, and the gradual onset of shortness of breath. People who carry the virus are the source of infection even during the incubation period. Early detection of the disease or the virus carrier is the key to prevent further spread. However, confirmation of the infection requires Nucleic Acid Detection Kit. The virus can be identified in swabs, secretions, and sputum from the respiratory tract, blood, or feces (7).

Computed tomography (CT) scan is highly important in the diagnosis and follow-up of lung disease treatment. In a review of different studies, one may find that the imaging features of COVID-19 pneumonia are varied; from their natural appearance to diffuse changes in the lungs. In addition, different radiological patterns are observed at different times over the course of the disease. Since the onset of symptoms and acute respiratory distress syndrome (ARDS) was short- lived in first cases with COVID-19 pneumonia, early detection of the disease is essential for the management of these patients (8).

Numerous studies have been performed on the findings of CT scans in COVID-19 patients and the results are inconsistent (3, 8-46). In a systematic review and meta-analysis, a structured review of all documentation and their composition can provide a more comprehensive picture of all dimensions of the subject. One of the main goals of meta-analysis, which is a combination of different studies, is to reduce the differences between parameters by increasing the number of studies involved in the analysis process. Another noteworthy goal of meta-analysis is to find the inconsistencies between the results and their causes (47-49). The present meta-analysis was performed to evaluate CT findings in COVID-19 patients at the time of admission.

## 2. Method

### 2.1. Study protocol

All research steps were taken according to the Meta-analyses Of Observational Studies in Epidemiology (MOOSE) protocol (49) and the final report was based on the Preferred Reporting Items for Systematic Reviews and Meta-Analyses (PRISMA) guideline (Supplementary file [SF]1)(64). Each stage of the study was conducted by two independent authors. Disagreements were resolved by discussion or a third author was involved. We registered this review at PROSPERO (registration number: CRD42020178078) (SF2). Available at: https://www.crd.york.ac.uk/prospero/display_record.php?RecordID=178078.

### 2.2. Literature search

We searched the Web of Science (ISI), Ovid, Science Direct, Scopus, EMBASE, PubMed/Medline, Cochrane Library (Cochrane Database of Systematic Reviews - CDSR), EBSCO, CINAHL and Google scholar databases using the following keywords: “2019 nCoV”, “Novel coronavirus”,”COVID-19”, “Novel coronavirus 2019”, “Wuhan pneumonia”, “Wuhan coronavirus”, “acute respiratory infection”, “COVID-19”, and “SARS-CoV-2”, “CT scan”, “Computed tomography”, “Radiology”, “Radiography”, “Clinical Characteristics”, “clinical features”, and “COVID-19”. An example of a combined search within PubMed is as follows: (“2019 nCoV”, OR “Novel coronavirus”, OR “COVID-19”, OR “Novel coronavirus 2019”, OR “Wuhan pneumonia”, OR “Wuhan coronavirus”, OR “acute respiratory infection”, OR “COVID- 19”, OR “SARS-CoV-2”) AND (“CT scan” OR “Computed tomography” OR “Radiology” OR “Radiography” OR “Clinical characteristics” OR “clinical features” OR “COVID-19”).

The search was conducted on March 20, 2020. Additional studies were identified by reviewing the reference lists of relevant articles. No language restrictions were applied. Since the present study was based on a regular review of previous studies, approval of the organizational review board and patient satisfaction was not necessary. The research received no specific funding. Grey literature was found at medrxiv (https://www.medrxiv.org/) and manual search of related articles was also conducted.

### 2.3. Inclusion and exclusion criteria

Inclusion criteria were all cross-sectional epidemiological studies aimed at examining chest CT scan findings in COVID-19 patients from January 1, 2020 until March 20, 2020 without language restrictions. Exclusion criteria were as follows: 1) non-random sampling, 2) duplicate studies, 3) studies on non-adult population (more than 10% of sample size being children), 5) being irrelevant, 6) sample size smaller than 10 participants, 7) diagnostic intervention for COVID-19 other than laboratory confirmation, 8) CT scans findings have not been verified by at least one radiology expert 9) poor quality in qualitative evaluation, and 10) case reports, review articles, and letters to the editor without quantitative data.

### 2.4. Study selection and data extraction

Two authors independently presented the results of the initial search with the title and abstract. At this stage, duplicate and unrelated studies were excluded. Duplicate studies were identified manually or using EndNote X9. Both authors then reviewed the full text of appropriate articles for the inclusion and exclusion criteria. Finally, the authors independently extracted the data from the articles. Any discrepancies between the data extractors were resolved by consensus or by a third author. It should be noted that when an article reported duplicate information from the same patients, both reports were combined to obtain the most complete data, but was considered as one case.

Data summary form includes the following items: First author’s name and year of publication, country and province, article references, study design, mean age and standard deviation, average duration from onset of symptoms until admission, time of performing CT scan, COVID-19 detection method, patient description, sample (respiratory secretions, blood, etc.), sample location (nasal, pharyngeal, etc.), number of patients (total, male and female), number of patients referred to the intensive care unit (ICU), quality of articles, positive chest CT scan in COVID-19 patients, number of positive chest CT scan findings in COVID-19 patients available in the studies.

### 2.5. Qualitative evaluation

Based on the type of studies, the adapted Newcastle–Ottawa Scale (NOS) was used to evaluate the risk of bias (65). Three categories were defined: studies with scores less than 6 were low quality studies, studies with scores 6 or 7 were medium quality, and studies with scores 8 or 9 were high quality studies.

### 2.6. Statistical analysis

I^2^ index (with values ranging from 0 to 100%) was used to evaluate the heterogeneity between studies; values above 75% indicate high heterogeneity, 50-74% indicate significant heterogeneity, 25-49% indicate moderate heterogeneity, and values below 25% indicate low heterogeneity (66, 67). In addition, P< 0.1 was also defined for heterogeneity. Meta-analysis was performed with at least three studies. In case of low heterogeneity, the fixed effects model and otherwise, the random effects model was used to combine the studies. Results were reported as pooled prevalence and 95% CI. To find the cause of heterogeneity, we could not perform subgroup analysis or meta-regression due to limitations. Sensitivity analysis for meta-analyses with at least five studies was performed by omitting one study at a time to evaluate the consistency of the results. Funnel plots and the Begger’s and Egger’s test were used to evaluate publication bias. All analyses were performed using Comprehensive Meta-Analysis ver.2. P-values less than 0.05 were considered statistically significant.

## 3. Results

### 3.1. Description of included studies

We identified 2264 potential articles from databases. After removing duplicates, there were 766 articles left. After evaluating the titles and abstracts, 85 articles were removed for at least one of the following reasons: non-random sampling (N=10), studies on non-adult population (N=3), being irrelevant (N=650), diagnostic intervention for COVID-19 other than laboratory confirmation (N=3), CT scans findings have not been verified by at least one radiology expert (N=2), poor quality in qualitative evaluation (N=0), and case reports, review articles, and letters to the editor without quantitative data (N=58). Finally, 40 eligible studies with 4183 patients with COVID-19 were used for meta-analysis. This process is illustrated in the SF3. The mean age of the study participants was 50.52 years (95% CI: 50.87-52.17).

### 3.2. Positive chest CT findings for COVID-19

The rate of positive chest CT scan in patients with COVID-19 was 94.5% (95% CI: 91.7-96.3). The lowest and highest estimates were for studies by Azami (61.5%) and many other studies (100%), respectively (Figure 1).

**Figure 1:**
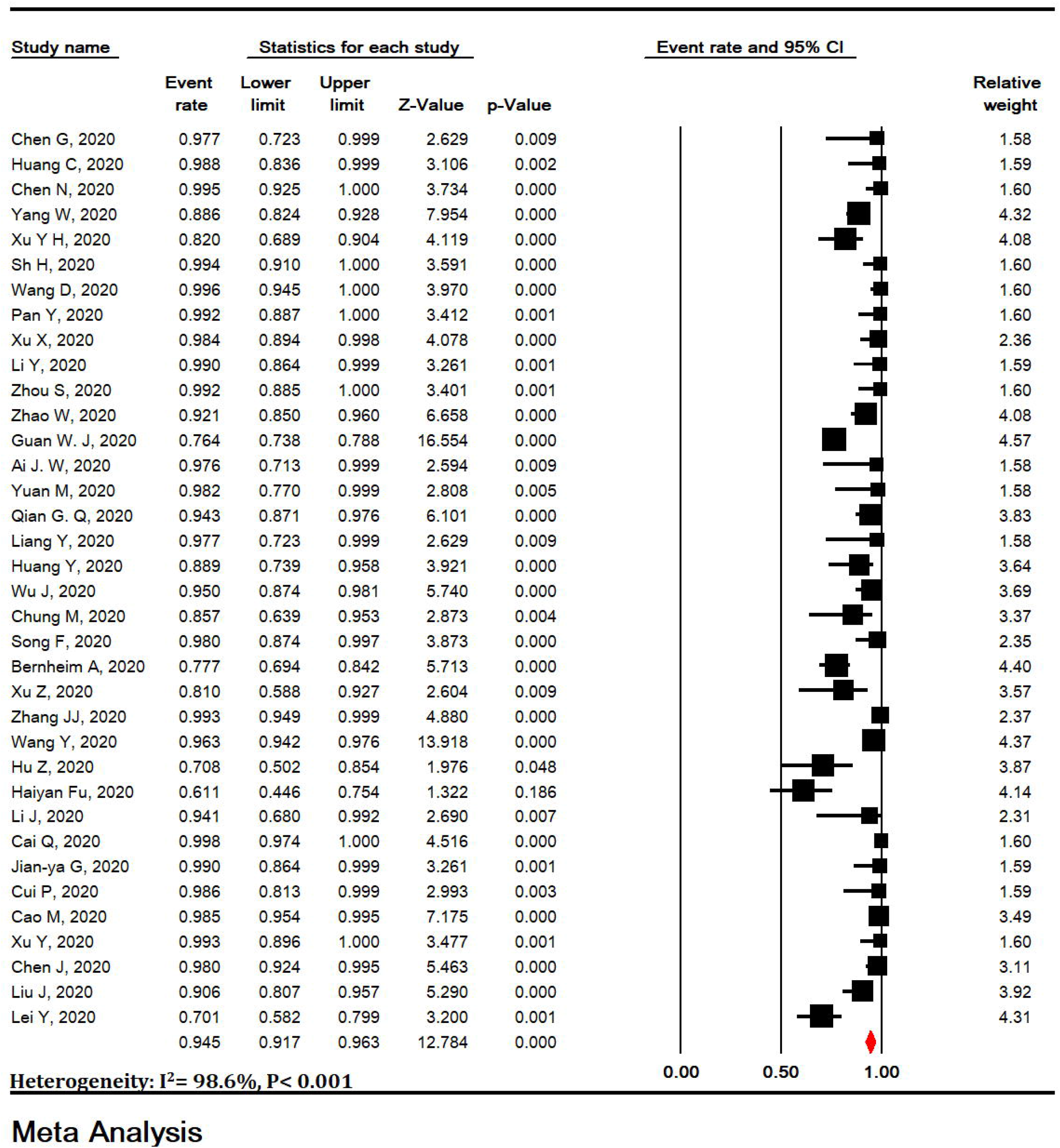
Meta-analysis of positive chest CT scan in patients with COVID-19.

### 3.3. Bilateral lung involvement

Bilateral lung involvement in chest CT scan of patients with COVID-19 pneumonia was estimated to be 79.1% (95% CI: 70.8-85.5) (Figure 2).

**Figure 2:**
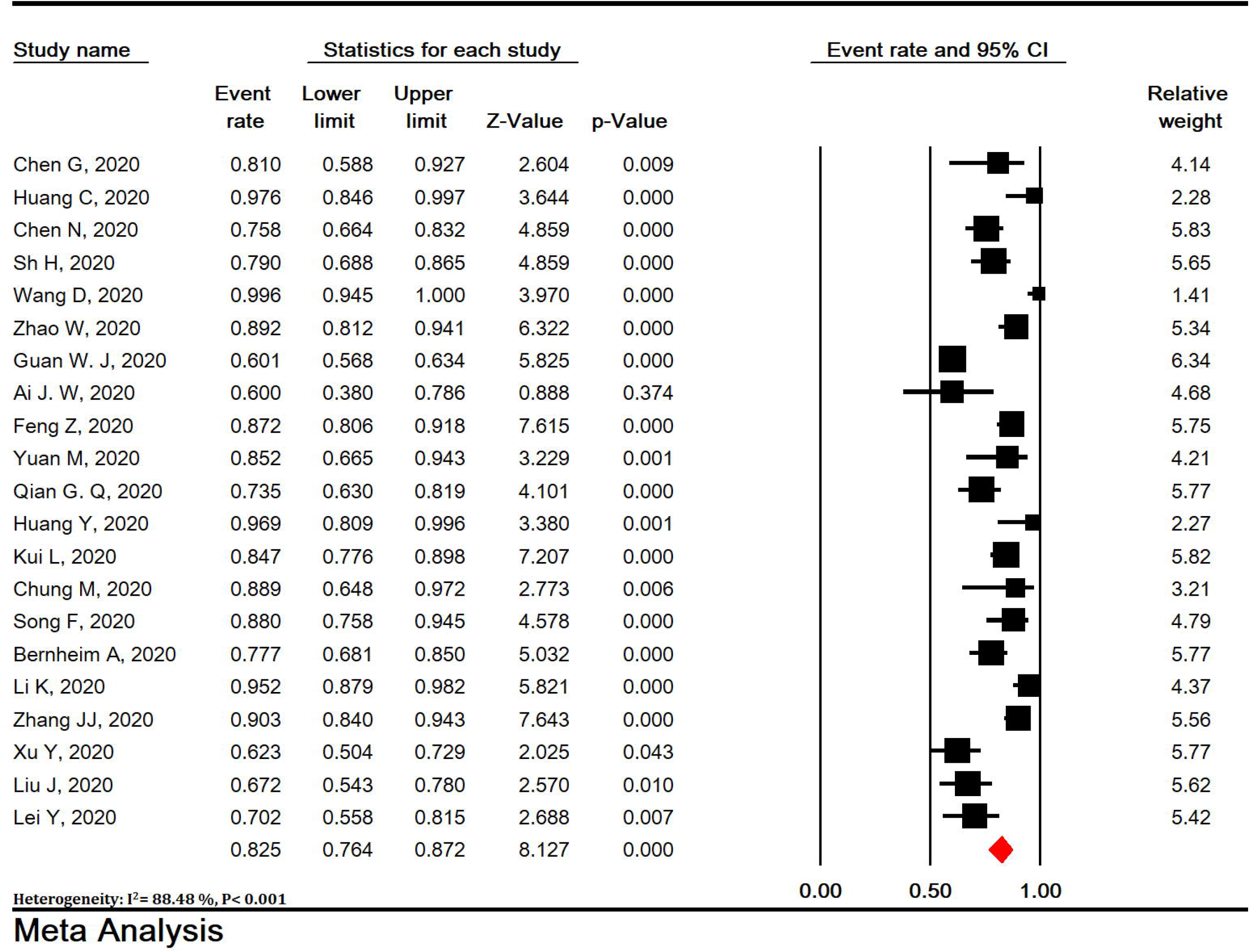
Meta-analysis of bilateral lung involvement in chest CT scan of patients with COVID- 19 pneumonia.

### 3.4. Predominant chest CT scan patterns

Pure ground-glass opacity (GGO), mixed (GGO pulse consolidation or reticular), consolidation, reticular, and presence of nodule findings in chest CT scan of COVID-19 pneumonia patients were respectively estimated to be 64.9% (95% CI: 54.1-74.4), 49.2% (95% CI: 35.7-62.8), 30.3% (95% CI: 19.6-43.6), 17.0% (95% CI: 3.9-50.9) and 16.6% (95% CI: 13.6-20.2) (Figure 3).

**Figure 3:**
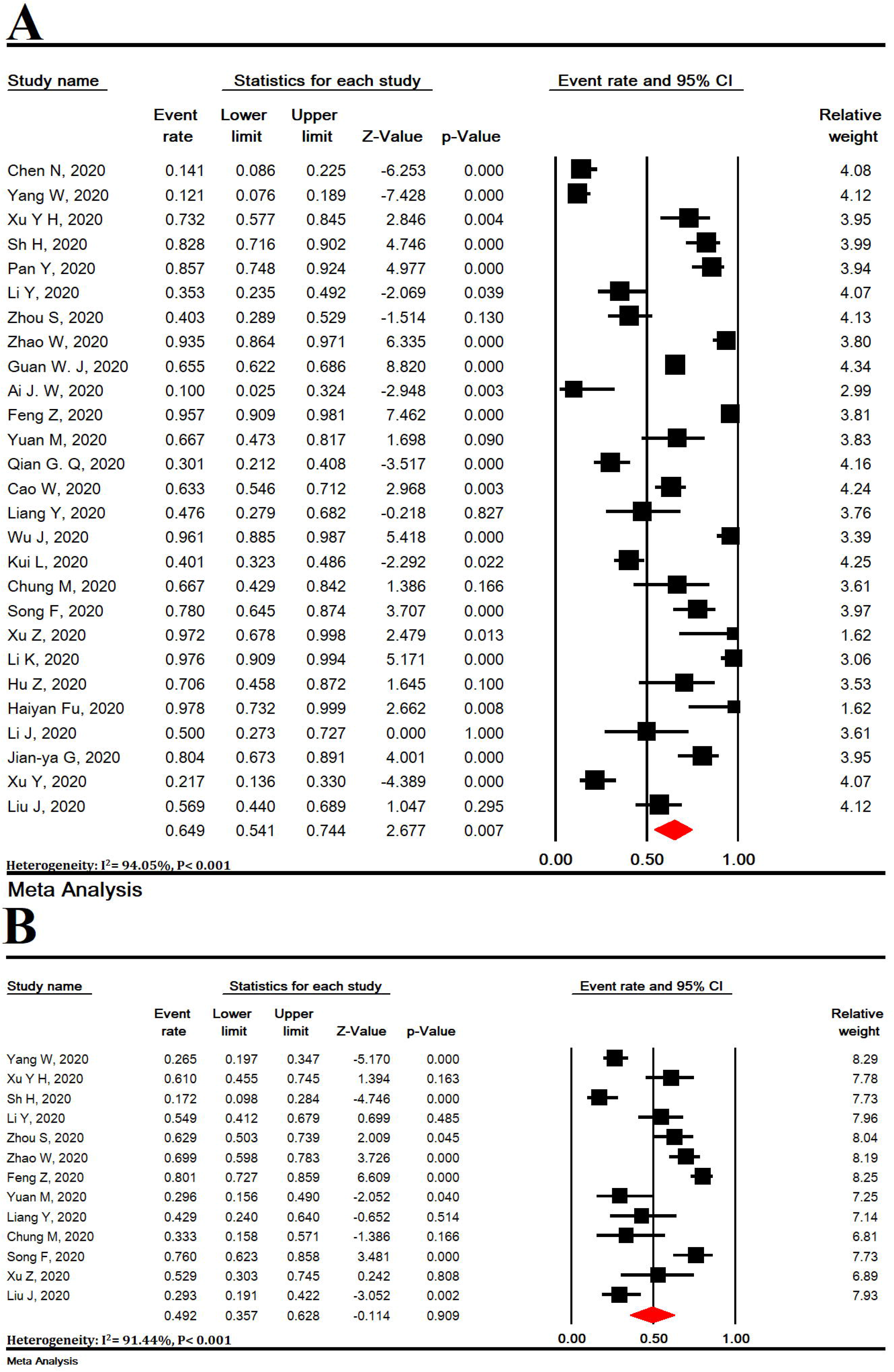
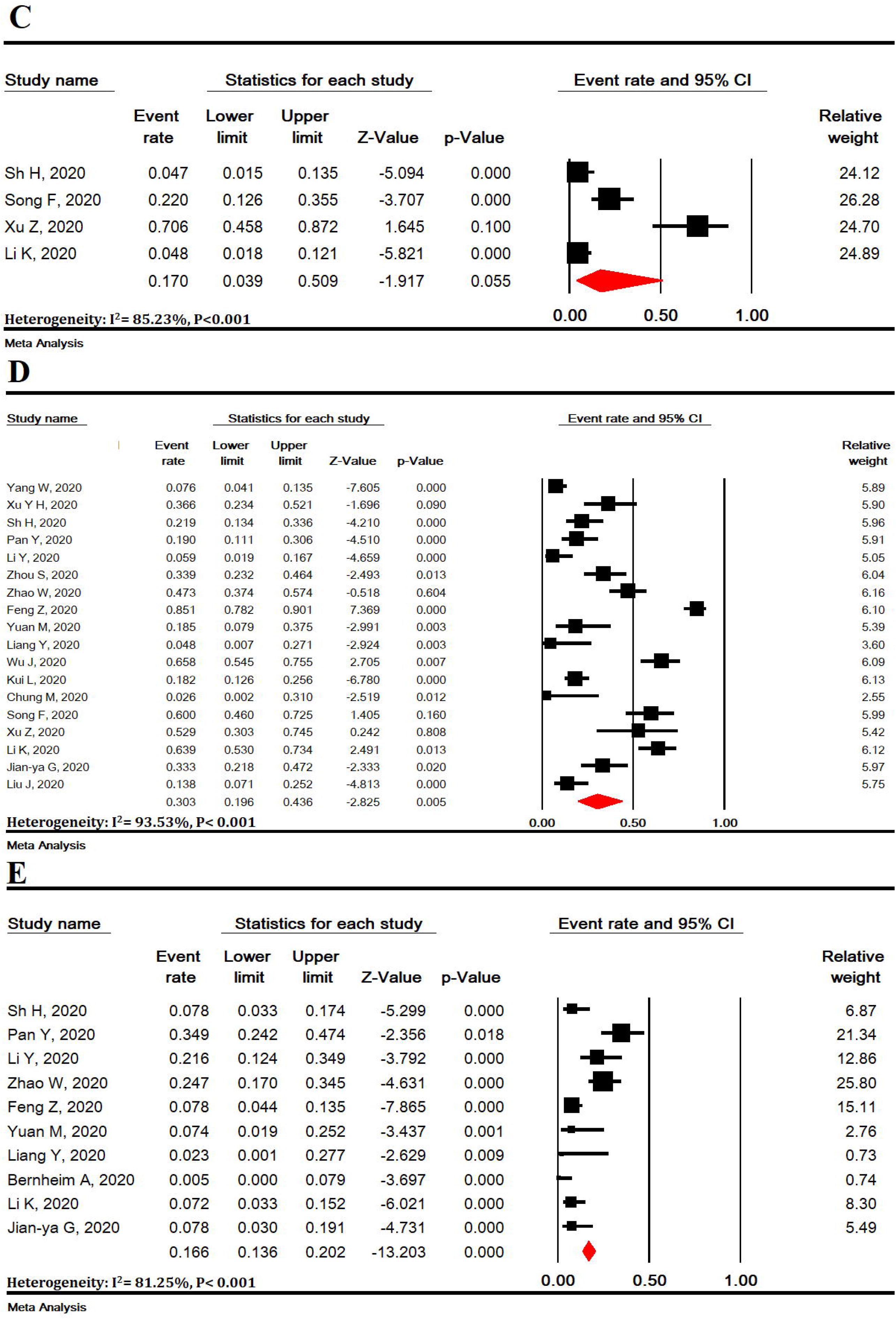
Meta-analysis of pure ground-glass opacity (GGO) (A), mixed (GGO pulse consolidation or reticular) (B), consolidation (C), reticular (D), and presence of nodule (E)€ findings in chest CT scan of COVID-19 pneumonia.

### 3.5. Other chest CT scan features

Other chest CT scan features are shown in Figure 4 and thickened interlobular septa was 63.6% (95% CI: 52.1-73.8), vascular enlargement was 61.4% (95% CI: 40.4-79.0), air bronchogram sign was 53.5% (95% CI: 40.3-66.2), bronchial wall thickening was 19.8% (95% CI: 12.6-29.6), bronchiolectasis was 19.9% (95% CI: 6.5-47.2), fibrous stripes was 17.2% (95% CI: 5.2-44.2), crazypaving pattern was 21.7% (95% CI: 13.8-32.5), thickening of the adjacent pleura was 30.0% (95% CI: 16.1-48.8), pleural effusion was 6.9% (95% CI: 4.7-10.1) and lymphadenopathy was 4.7% (95% CI: 3.0-7.5) (Figure 4).

**Figure 4:**
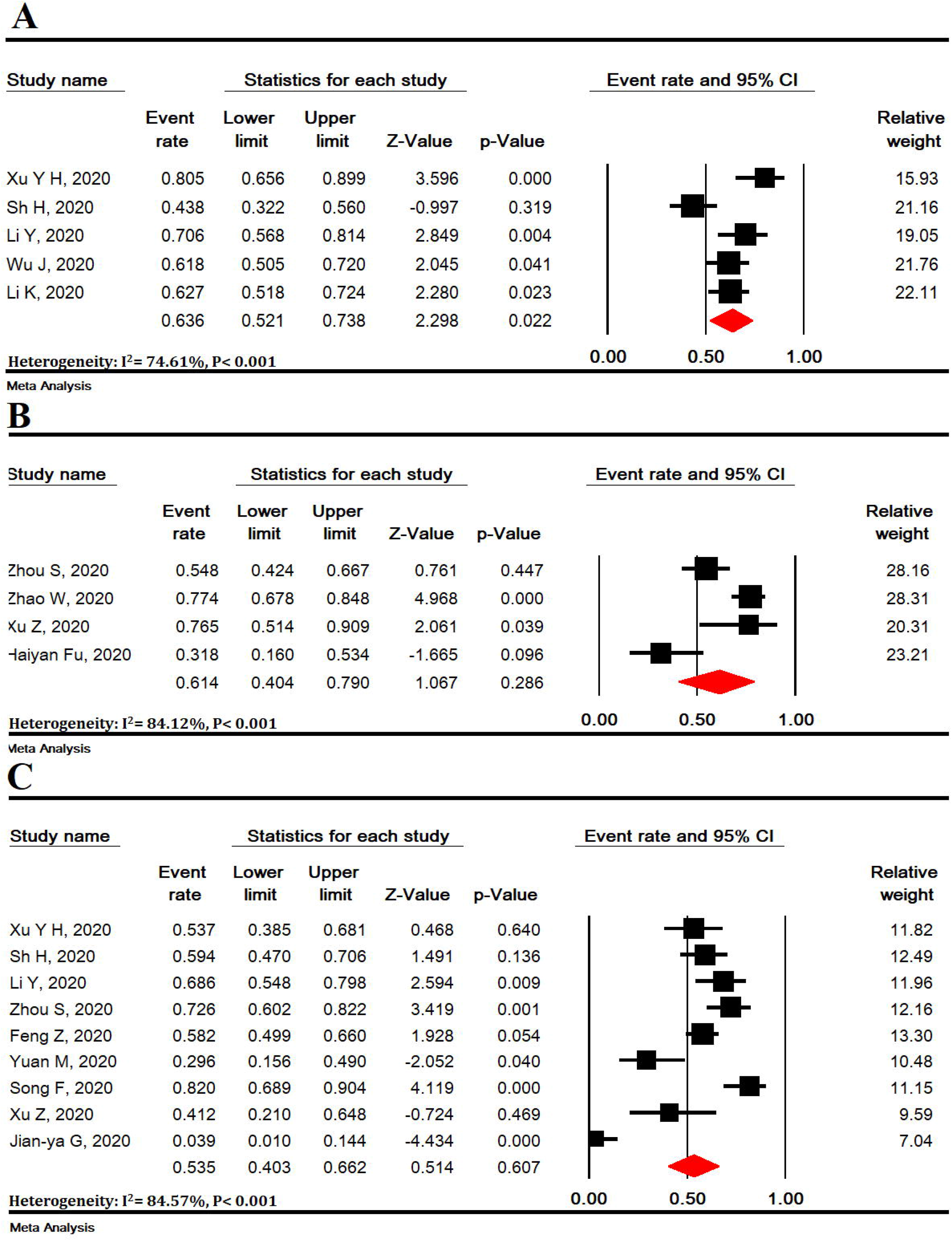
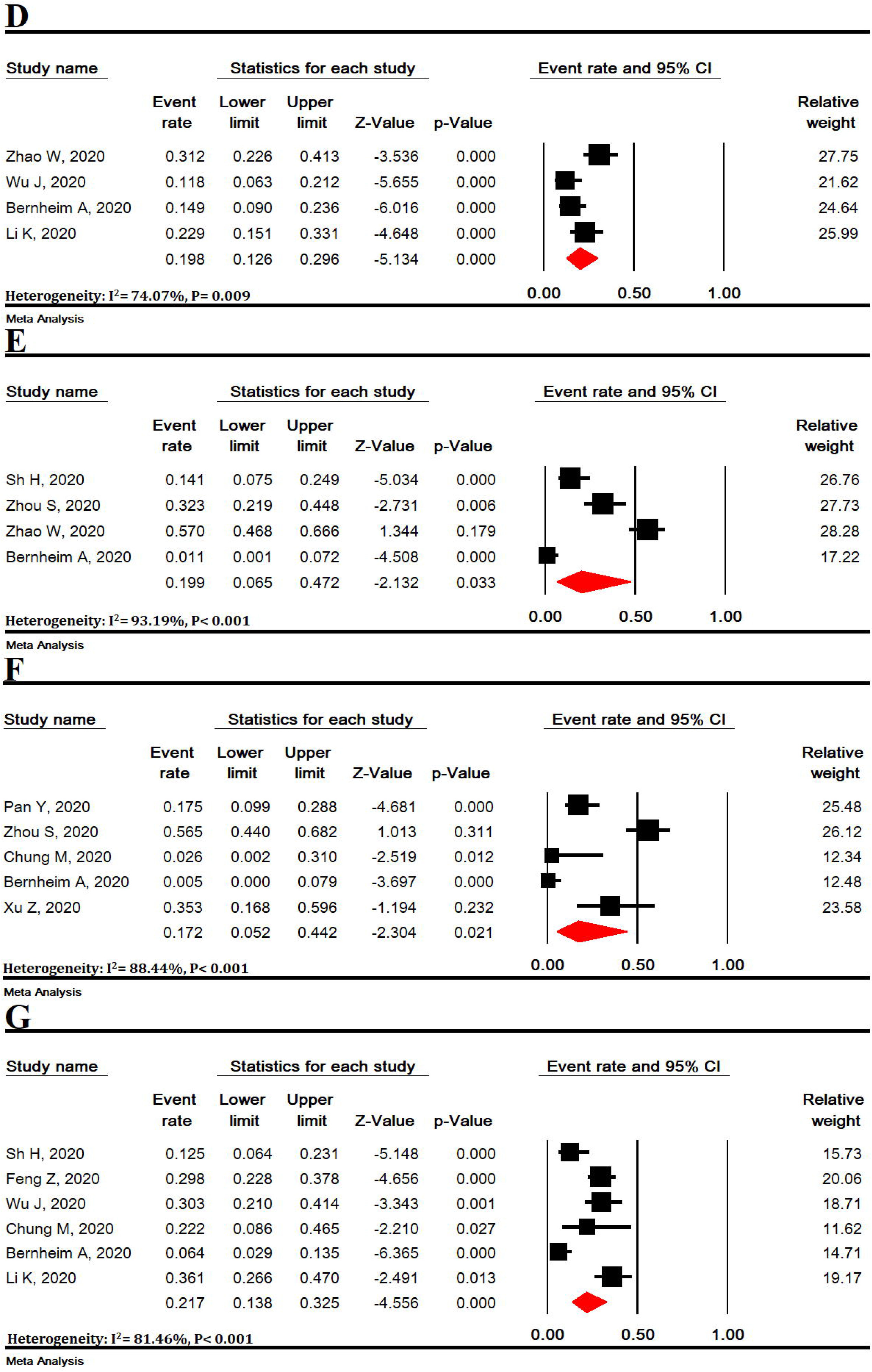
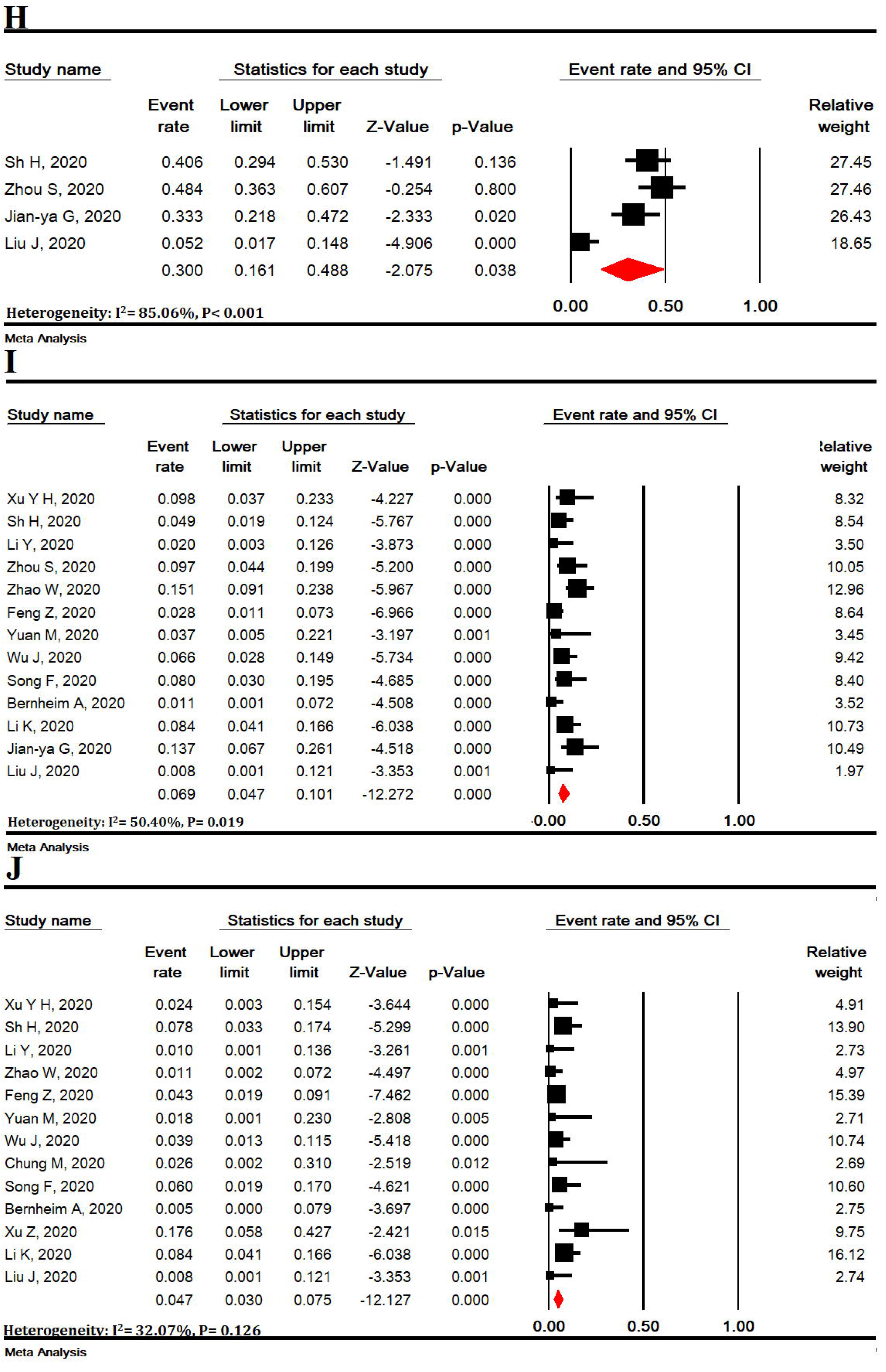
Meta-analysis of thickened interlobular septa (A), vascular enlargement was (B), air bronchogram sign (C), bronchial wall thickening was (D), bronchiolectasis (E), fibrous stripes (F), crazypaving pattern (G), thickening of the adjacent pleura (H), pleural effusion (I) and lymphadenopathy (J) findings in chest CT scan of COVID-19 pneumonia.

### 3.6. Lesions distribution

The distribution of lung lesions in patients with COVID-19 pneumonia was as follows: peripheral (70.0% [95% CI: 57.8-79.9]), central (3.9% [95% CI: 1.4-10.6]), and peripheral and central (31.1% [95% CI: 19.5-45.8]) (SF4).

### 3.7. Lobes involvement

Pulmonary lobes involvement in patients with COVID-19 pneumonia was as follows (Figure 5): right upper lobe (58.4% [95% CI: 33.6-79.5]), right middle lobe (49.7% [95% CI: 23.0-76.6]), right lower lobe (86.5% [95% CI: 57.7-96.8]), left upper lobe (64.5% [95% CI: 37.3-84.7]), and left lower lobe (81.0% [95% CI: 50.5-94.7]).

**Figure 5:**
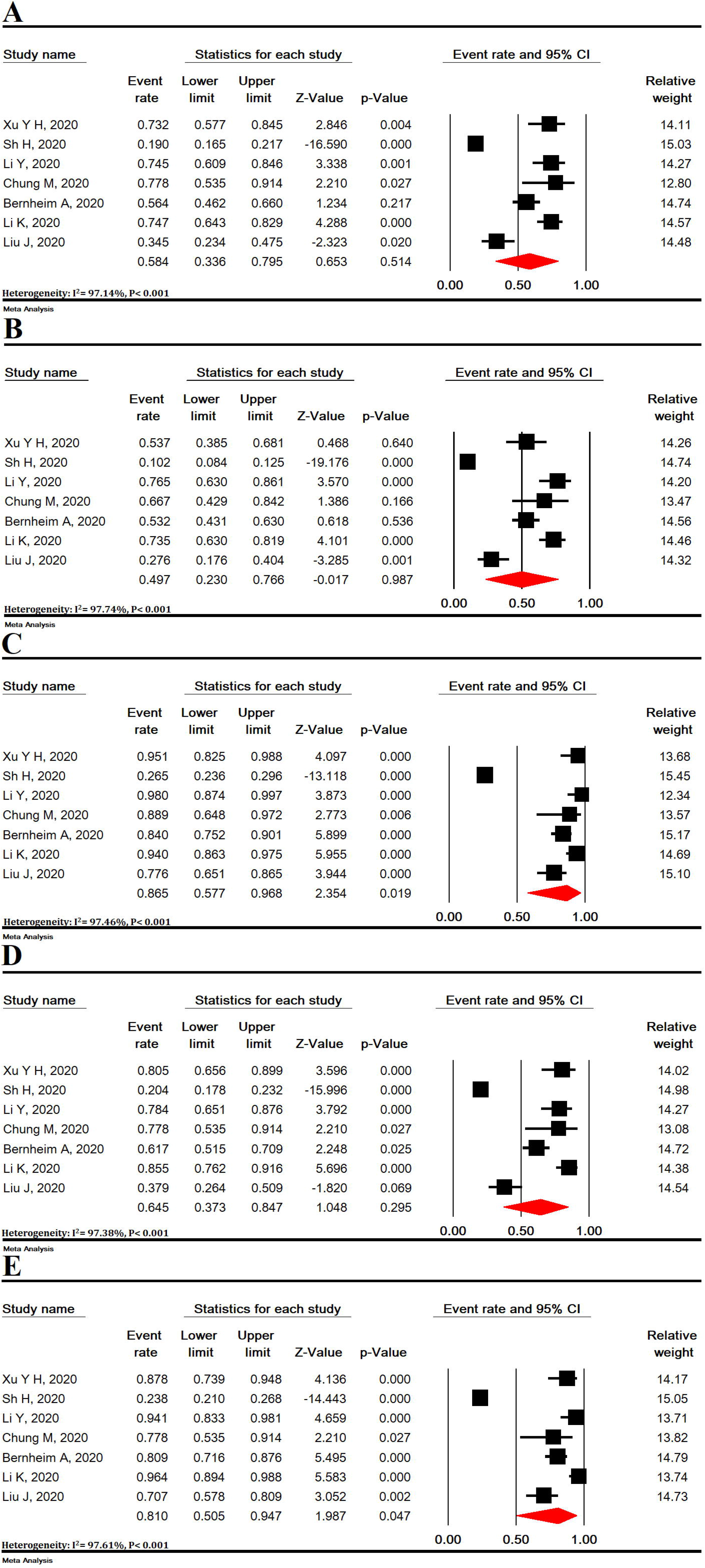
Meta-analysis of right upper lobe (A), right middle lobe (B), right lower lobe (C), left upper lobe (D), and left lower lobe (E) involvement in chest CT scan of COVID-19 pneumonia.

### 3.8. Number of involved lobes

In patients with COVID-19 pneumonia, the number of involved lobes was as follows (SF5): One lobe (58.4% [95% CI: 33.6-79.5]), two lobes (31.1% [95% CI: 19.5-45.8]), three lobes (10.5% [95% CI: 8.0-13.8]), four lobes (20.1% [95% CI: 15.4-25.9]), and five lobes (43.2% [95% CI: 34.2-52.6]).

### 3.9. Sensitivity analysis

Sensitivity analysis was performed for all meta-analyses and it showed that the overall result remains robust after the omission of one study at a time (SF6-10).

### 3.10. Publication bias

Publication bias was evaluated for studies that showed positive chest CT scan of COVID-19 patients (Begg’s test= 0.774 and Egger’s test< 0.001) and for studies that showed bilateral lung involvement in chest CT scan of COVID-19 patients pneumonia (Begg’s test= 0.194 and Egger’s test< 0.001) (SF11).

## 4. Discussion

The present study is the first systematic review and meta-analysis that extensively examines chest CT scan findings in COVID-19 patients. COVID-19 is the outbreak of a new disease with serious consequences for public health. Chest CT scan is an important part of disease detection for patients suspected of having COVID-19 infection and may help early detection of lung malformations for the purpose of screening highly suspected patients, especially those with negative initial RT-PCR screening (50). In fact, given the limited number of RT-PCR kits in many centers and the likelihood of false negative RT-PCR results, the National Health Commission of China has encouraged clinical findings and chest CT scan (51). Our review showed some imaging findings that are often seen in patients with COVID-19. The present study showed that 94.5% of COVID-19 patients had positive chest CT scan findings, while bilateral lung involvement in chest CT scan of COVID-19 pneumonia patients was 79.1%. It is important to observe the high incidence of bilateral organizing pneumonia in these patients. This suggests that corticosteroids may be an option to suppress this immune response in lung parenchyma of COVID-19 pneumonia.

Viruses are a common cause of respiratory tract infection. Imaging findings of viral pneumonia are varied, and may overlap with other infectious and inflammatory lung diseases. Viruses in the same viral family have a similar pathogenesis, so chest CT scan may help identify distinct patterns and features in immunocompromised patients (52). Meta-analysis of initial data suggests that chest CT scan findings for 2019-nCoV have many features similar to other viruses such as the Middle East Respiratory Syndrome (MERS-CoV) and Severe Acute Respiratory Syndrome (SARS-CoV) (53).

In the present study, most pulmonary lesions include bilateral lung involvement with multiple lung lobes (predominantly right lower lobe and left lower lobe), with dominant distribution in the peripheral portion of the lungs. Studies have shown that influenza pneumonia tends to affect the lower lobes (52, 54). Wang et al. also showed that H7N9 pneumonia has a predominant distribution in the right lower lobe (54). Both H1N1 and SARS pneumonia are more peripherally distributed (55, 56), whereas no lobe infection is found in H5N1 influenza (57). However, lung involvement with peripheral predominance has also been observed in SARS and MERS. Similarly, previous coronavirus pneumonias have a similar pattern. The dominant peripheral distribution for COVID-19 was shown in our study. Such a distribution is obvious at first glance. This feature of chest CT scan is caused by alveolar injury and pulmonary interstitial edema. We also observed some chest CT scan features of COVID-19 that differ from chest CT scan features of SARS and MERS. Unifocal involvement is more common than multifocal involvement in patients with SARS and patients with MERS (58, 59). However, contrary to what is seen in chest CT scan of patients with COVID-19, multi-lobe involvement was more common than single-lobe involvement in the present meta-analysis. Thus, more than two lobes are likely to be involved in this disease. To the best of our knowledge, these findings have not been reported in the literature related to SARS and MERS.

Our results showed that the most common findings of imaging were pure GGO, GGO with mixed consolidation or reticular pattern, interlobular septal thickening, and consolidation. Reticular and nodular pattern were relatively small, which may be explained in the first stage of the disease. In H7N9 pneumonia, most cases showed consolidation (58). Each of the chest CT scan patterns in our patients is nonspecific and may overlap with other microorganism infections such as H7N9 pneumonia, H1N1 virus infection, SARS, MERS, and avian influenza A (H5N1) (56-59).

GGO, consolidation, and interlobular septal thickening are the most common chest CT scan findings of H1N1 influenza pneumonia, too (55). Based on the present meta-analysis, pure GGO is a common finding of about 65%, and 49% GGO with interlobular septal thickening/consolidation in COVID-19 pneumonia patients. Thus, these features of chest CT scan can be seen in most patients. This finding, along with the dominant distribution in the peripheral part of the lungs, is not common in other viral pneumonia (8, 18, 27, 29, 56, 57).

In addition, we found that all features of chest CT scan that exist in the initial chest CT scan of patients with COVID-19, like GGO and consolidation, and other chest CT scan features such as vascular enlargement, interlobular septal thickening, and air bronchogram sign are also present in the chest CT scan of SARS and MERS. The low incidence of pleural effusion and lymphadenopathy noted in our data was also a feature of chest CT scan in previous studies about SARS (53). This may be due to the inherent anatomical features of the lower lobe bronchus. The right lower lobe bronchus is tighter than other bronchi of the lung, and the angle between the right lower lobe and the long axis of the trachea is smaller, so it is more viral at early stages. Most likely, it attacks the bronchial branches of the lower lobe and causes infection.

Interestingly enough, we found that most patients have vascular enlargement lesions (61.4%) that may be caused by an acute inflammatory response. However, vascular changes are not similar to changes in malignant lesions such as lung adenocarcinoma that cause vascular dilatation or irregularity and vascular convergence, which may be due to chronic progression and tumor infiltration (17, 60).

Angiotensin II converting enzyme is a key molecule involved in the development and progression of acute lung failure. COVID-19 induces direct lung injury by involving angiotensin converting enzyme, which contributes to the progression of alveolar injury (61). This may explain the pathological mechanism of GGO and consolidation as well as rapid changes in chest CT scan findings. Our results support the observed process, according to which, bilateral GGOs or mixed GGOs in chest CT scan should prompt the radiologist to recommend COVID-19 as a possible diagnosis (62, 63).

In summary, this work is a meta-analysis on preliminary studies of chest CT scan findings about COVID-19, aimed at introducing the common imaging manifestations of the disease. Radiologists play an important role in the rapid identification and early detection of new cases, which can be useful not only for the patient but also for public health surveillance systems. It is important to recognize the fact that the appearance of CT findings about COVID-19 has some similarities to other viral diseases, especially those in the same viral family (SARS and MERS). Future studies are recommended to determine how CT scans of COVID-19 patients change after treatment.

## 5. Limitations

Limitations of this study include: 1. All studies were performed in China, and the severity of chest CT scan manifestations might be affected by ethnic factors; 2. Most patients were the hospitalized patients and patients with milder symptoms or those who were not hospitalized, which may cause bias in the results. 3. In most preliminary studies, chest CT scan findings were not separately reviewed according to patients admitted to the intensive care unit or the isolated ward, 4. Follow-up for chest CT scan during treatment until discharge was not included in our study, 5. Our results are based on CT findings during admission, but patients might have experienced symptoms before admission (because chest CT scan findings are influenced by the clinical course of the disease) and during this time, the patient might have received antiviral or antibacterial drugs, or steroid therapy, and this might have affected chest CT scan findings, 6. Since all studies were performed in 2020 in China with the same diagnostic method, we could not discover the cause of heterogeneity.

In conclusion, our study showed that chest CT scan has little weakness in diagnosis of COVID- 19 combined to personal history, clinical symptoms, and initial laboratory findings, and may therefore serve as a standard method for diagnosis of COVID-19 based on its features and transformation rule. Rapid detection may lead to early control of the transmission. By diagnosing viral pneumonia on CT scan, infected or suspected patients can be isolated and treated in a timely manner to optimize patient management, especially for hospitals or communities without Rt-PCR test kits. However, chest CT scan is still limited in terms of identifying specific viruses. It is important that radiologists recognize if chest CT scan findings for COVID-19 overlap with chest CT scan findings for diseases caused by different virus families, such as adenovirus. They should also recognize the differences and similarities with other viruses in the same family, such as SARS-CoV and MERS-CoV or other families such as influenza. According to the present meta-analysis, lesion distribution in patients with COVID-19 is likely to have peripheral distribution, bilateral involvement, lower lobe dominance, and multi-lobe distribution.

## Data Availability

Not applicable.

## Authors’ contributions

MA and MK acquired the data. MA and MK analyzed and interpreted the data. MA drafted the manuscript; MA and MK critically revised the manuscript for important intellectual content. MK supervised the study. All authors have read and approved the manuscript in its current state.

## Abbreviation

SARS-CoV-2: severe acute respiratory syndrome coronavirus-2
real-time PCR: real-time polymerase chain reaction
WHO: World Health Organization
COVID-19: Coronavirus Disease 2019
CT: computed tomography
MOOSE: Meta-analyses Of Observational Studies in Epidemiology
PRISMA: the Preferred Reporting Items for Systematic Reviews and Meta-Analyses
PROSPERO: The International Prospective Register of Systematic Reviews
ICU: intensive care unit
GGO: ground-glass opacity
MERS-CoV: Middle East Respiratory Syndrome
SARS-CoV: Severe Acute Respiratory Syndrome

## Supplementary files

Supplementary file 1: PRISMA checklist

Supplementary file 2: PROSPERO registration

Supplementary file 3: PRISMA flowchart

Supplementary file 4: Meta-analysis of the distribution of lung lesions in patients with COVID- 19 pneumonia was as follows: peripheral (A), central (B), and peripheral and central (C).

Supplementary file 5: Meta-analysis of one lobe involvement (A), two lobes involvement (B), three lobes involvement (C), four lobes involvement (D), and five lobes involvement (E).

Supplementary file 6: Sensitivity analysis of positive chest CT scan in patients with COVID-19.

Supplementary file 7: Sensitivity analysis of bilateral lung involvement in chest CT scan of patients with COVID-19 pneumonia.

Supplementary file 8: Sensitivity analysis of pure ground-glass opacity (GGO) (A), mixed (GGO pulse consolidation or reticular) (B), consolidation (C), reticular (D), and presence of nodule (E)€ findings in chest CT scan of COVID-19 pneumonia.

Supplementary file 9: Sensitivity analysis of thickened interlobular septa (A), vascular enlargement was (B), air bronchogram sign (C), bronchial wall thickening was (D), bronchiolectasis (E), fibrous stripes (F), crazypaving pattern (G), thickening of the adjacent pleura (H), pleural effusion (I) and lymphadenopathy (J) findings in chest CT scan of COVID-19 pneumonia.

Supplementary file 10: Sensitivity analysis of right upper lobe (A), right middle lobe (B), right lower lobe (C), left upper lobe (D), and left lower lobe (E) involvement in chest CT scan of COVID-19 pneumonia.

Supplementary file 11: Publication bias for positive chest CT scan of COVID-19 patients (A) and for studies that showed bilateral lung involvement in chest CT scan of COVID-19 patients pneumonia (B)

**Table 1:**
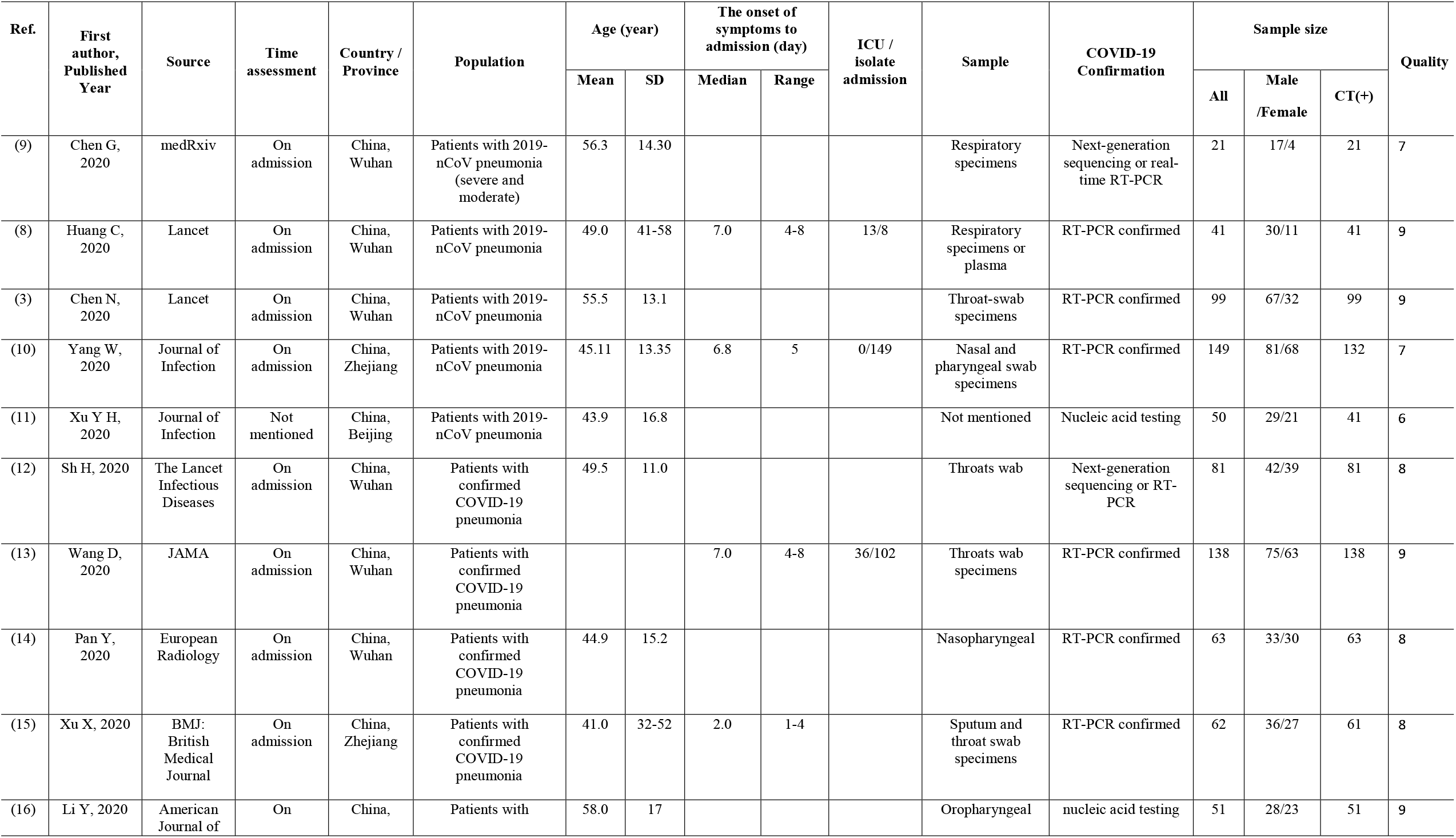
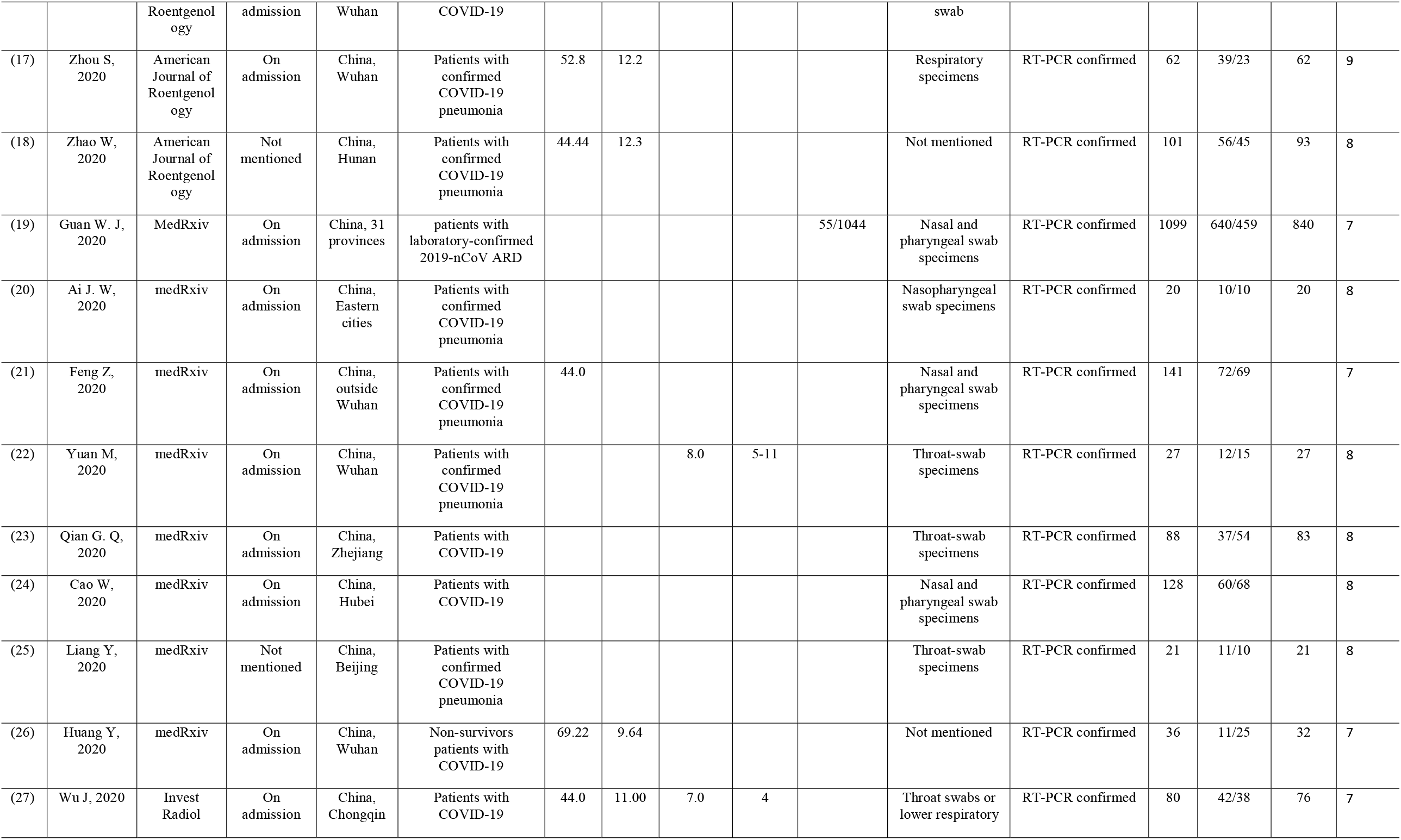
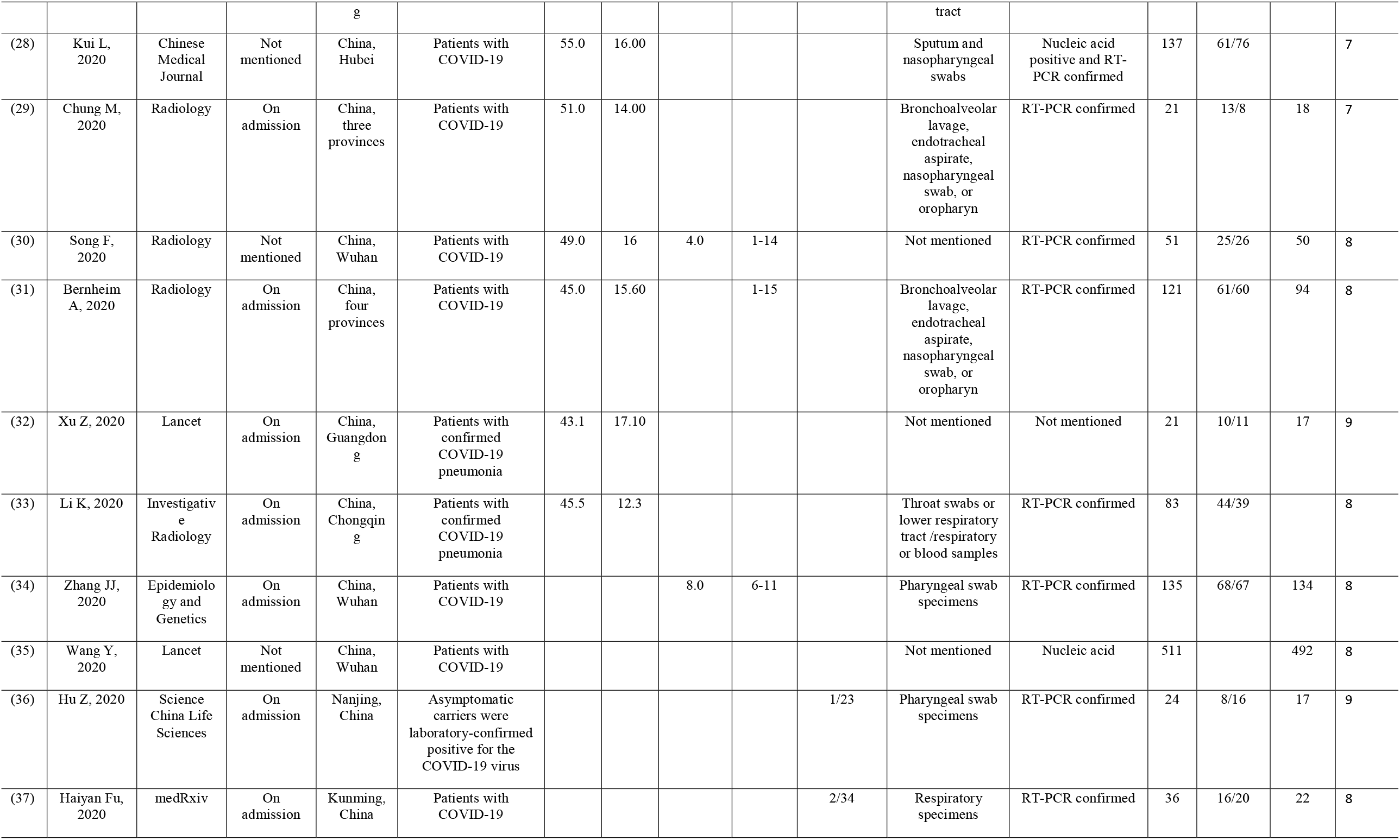
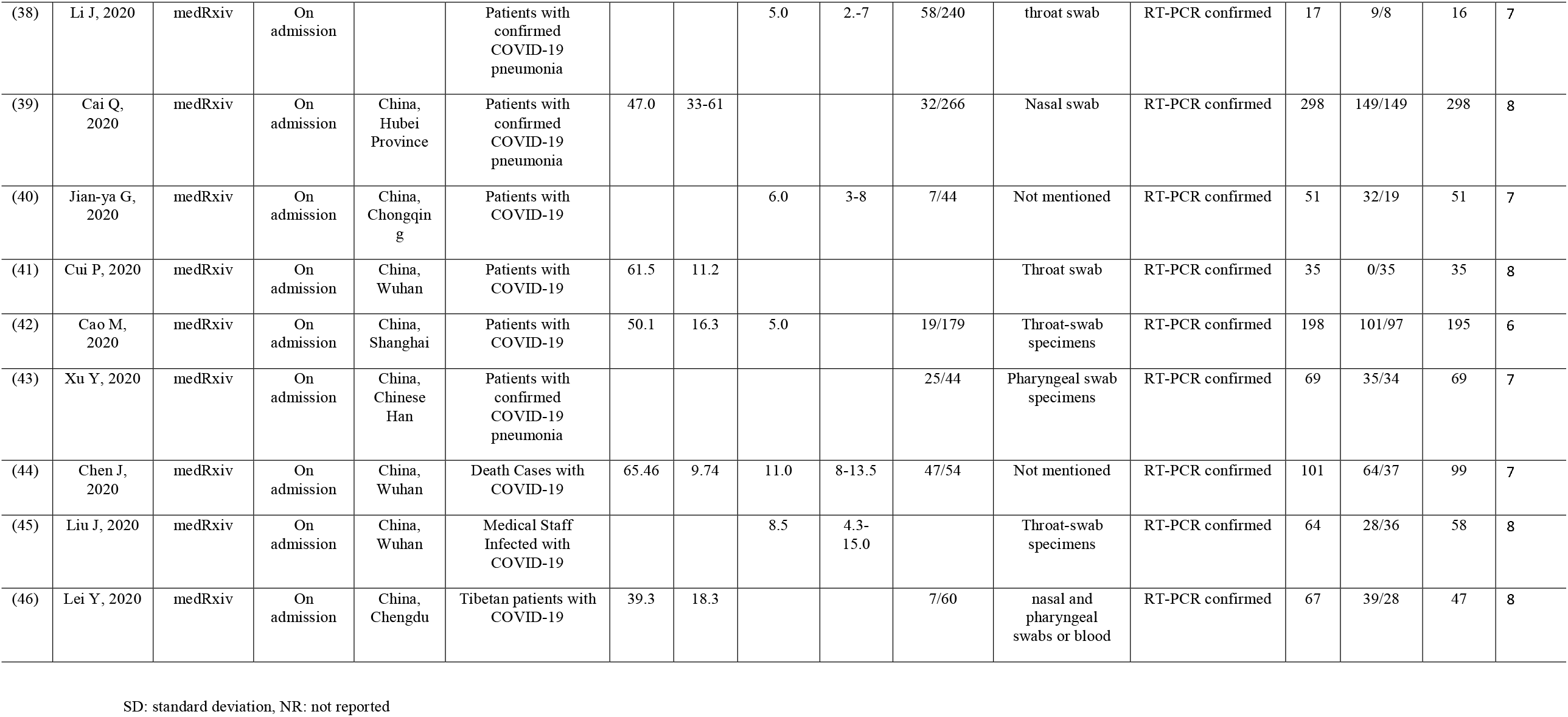
Characteristics of articles entered into meta-analysis

